# Genomic characterization of Dengue virus circulation in Ethiopia

**DOI:** 10.1101/2024.07.10.24310195

**Authors:** Adugna Abera, Houriiyah Tegally, Geremew Tasew, Eduan Wilkinson, Abraham Ali, Feyisa Regasa, Molalegne Bitew, Lucious Chabuka, Gaspary Mwanyika, Derek Tshiabuila, Jennifer Giandhari, Sureshnee Pillay, Jenicca Poogavanan, Monika Moir, CLIMADE Consortium, Moritz U.G. Kraemer, Kamran Khan, Carmen Huber, Getachew Tollera, Tobias F. Rinke de Wit, Cheryl Baxter, Richard Lessells, Dawit Wolday, Dereje Beyene, Tulio de Oliveira

## Abstract

In Ethiopia, dengue virus (DENV) infections have been reported in several regions, however, little is known about the circulating genetic diversity. Here, we conducted clinical surveillance for DENV during the 2023 nationwide outbreak and sequenced DENV whole genomes for the first time in Ethiopia. We enrolled patients at three sentinel hospital sites. Using RT-PCR, we screened serum samples for three arboviruses followed by serotyping and sequencing for DENV-positive samples (10.4% of samples). We detected two DENV serotypes (DENV1 and DENV3). Phylogenetic analysis identified one transmission cluster of DENV1 (genotype III major lineage A), and two clusters of DENV3 (genotype III major lineage B). The first showed close evolutionary relationship to the 2023 Italian outbreak and the second cluster to Indian isolates. Co-circulation of DENV1 and DENV3 in some regions of Ethiopia highlights the potential for severe dengue. Intensified surveillance and coordinated public health response are needed to address the threat of severe dengue outbreaks.

## Introduction

Dengue virus (DENV) is primarily transmitted by *Aedes* mosquitoes and causes significant epidemics in tropical and subtropical regions. Since the beginning of 2023, over 5 million cases and 5,000 dengue-related deaths have been reported in over 80 countries (1). In 2023, the WHO African region reported 171,991 suspected cases, 70,223 of which were confirmed cases with 753 fatalities. Outbreaks have been reported in 15 African countries: Burkina Faso has accounted for 85% of cases, followed by Ethiopia (8.2%), Mali (2.5%), and Côte d’Ivoire (2.2%) (2).

Ethiopia has been facing several outbreaks of dengue fever since 2013. The first recorded outbreak was reported in Dire Dawa City, which affected 9,441 individuals (3). A second outbreak occurred in Somale Godey Town and the Afar Region in 2014 and 2015, respectively (4). Since 2015, there has been an increase in the number of severe febrile illness cases in Godey, Dire Dawa town, and the Afar area, with no apparent cause. The first outbreak started in April 2023 in the Afar region, followed by Dire Dawa and the neighboring regions of Amhara and Oromia. There have been 27,577 reported cases and 21 deaths in 12 districts in the eastern part of Ethiopia (1,5).

DENV is a single-stranded positive-sense RNA virus, and its genome encodes three structural and seven nonstructural proteins. There are four serotypes of DENV, namely DENV1, DENV2, DENV3, and DENV4 (8, 9). When an individual is infected with one serotype, they develop immunity to that specific serotype for a long time. However, they only develop partial immunity to the other serotypes. In case of subsequent infections with a different serotype, the risk of severe dengue increases. Each serotype of the virus is further divided into several genotypes, which are categorized based on the envelope gene (E gene). However, different genotypes of the same serotype may vary in their ability to infect host cells and cause severe forms of the disease. Based on the genetic characterization, five genotypes have been defined for DENV1, six for DENV2, five for DENV3, and four for DENV4 (10–13). Recently, a nomenclature system has been proposed to further subdivide genotypes into major and minor lineages to aid global monitoring efforts (14).

Studies have shown that during dengue outbreaks, the emergence of new serotypes or changes in circulating genotypes of the virus in a particular region can lead to more severe outbreaks (15,16). Given that there are currently no approved medical countermeasures for treating severe dengue, and vaccines are currently not available to most countries, it is crucial to continuously monitor the circulating dengue genetic diversity in endemic areas. This information will be instrumental in developing and evaluating vaccines and treatments (17), and in responding effectively to dengue outbreaks.

Although there have been multiple outbreaks and geographical expansion of DENV in Ethiopia, there is currently no documented characterization of the circulating diversity of DENV, and no publicly available whole genome sequences. In this study, we address this gap by identifying circulating DENV serotypes and genotypes among individuals with febrile illness in three hospital-based sentinel sites in Ethiopia and conduct phylogenetic analysis to determine spatial and temporal patterns of dengue transmission in Ethiopia.

## Materials and methods

### Patient characteristics and study settings

We conducted a cross-sectional hospital-based study in Afar, Gambela, and Dire Dawa city administration (Supplementary Figure 1). Between December 2022 and November 2023, we enrolled outpatients and inpatients presenting with fever (>37.5°C) at three major public hospitals in Dubti (Afar region), Dilchora (Dire Dawa), and Gambela. These hospitals serve as sentinel sites for acute febrile illnesses in the country and were selected due to their previous history of arbovirus outbreaks and serological evidence for arboviruses (5, 18).

### Recruitment of participants

We obtained informed consent from all participants aged 18 years and older. For individuals aged 1 to 17 years, we obtained informed consent from a parent or guardian and, where appropriate, assent from the child.

### Eligibility/exclusion criteria

The inclusion criteria for the study were: a fever of more than 37.5 °C, following the 2009 WHO Dengue classification (19), joint pain, rash, headache, myalgia, retro-ocular discomfort, stomach pain, and hemorrhagic presentation.

Patients with severe, and established chronic clinical illnesses such as people living with HIV, malignancies, and known metabolic disorders were excluded from the study.

### Sample collection and processing

Each participant provided five milliliters of whole blood, which was collected into serum separator tubes. At the Afar site, blood sample collection took place between April 25, 2023, and May 15, 2023. The serum was isolated and collected in tubes containing 1 ml DNA/RNA shield and stored at −20 °C during field collections. It was then transported in a cold chain to the laboratory of the Ethiopian Public Health Institute (EPHI) and stored at −20 °C until analysis for PCR and sequencing. Blood samples from Dire Dawa and Gambela sites were collected between June 2023 and October 2023, stored at −20 °C, and then shipped in a cold chain to the laboratory of the EPHI, where they were stored at −80 °C until analysis.

### RNA extraction and PCR amplification

We performed RNA extraction within five days of receiving the serum samples at the EPHI laboratory from each site. The RNA extraction was carried out using a Bioer NPA-32P instrument (Bior Technology, Zhejiang, China), which is a high-throughput automated workstation. We used the MagaBio Plus RNA Purification (Bior Technology, Zhejiang, China) for the extraction process, following the manufacturer’s instructions. Briefly, we prepared 96-well plates and added 300 µl of thawed and briefly vortexed serum samples to columns #1 and #7. The plate was then placed into the instrument, which agitated the plates every 10 seconds for 9 minutes until the extraction was completed, resulting in 70 µl RNA extracts.

The remaining serum samples from the Afar region were stored at −20°C for later whole genome sequencing. Meanwhile, samples from the Dire Dawa and Gambela sites were stored at −80°C. In February 2024, some of the qualifying serum samples were sent to the CERI and KRISP laboratories in South Africa for whole genome sequencing.

On the same day that we extracted the RNA, we used the CDC Trioplex real-time RT-PCR assay to screen for arboviral infections such as DENV, CHIKV, and ZIKV. This assay includes a set of published oligonucleotide primers and dual-labeled hydrolysis Taqman probes. To run the test, we combined 10 µl of sample RNA with 12.5 µl of PCR master mix reaction buffer (SuperScript™ III One-Step RT-PCR System with Platinum™ Taq DNA Polymerase, ThermoFischer Scientific), 1 µM virus-specific primers, 0.3 µM dengue-specific probe, 0.15 µM chikungunya-specific probe, 0.15 µM Zika virus-specific probe, and nuclease-free water in a 96-well optical PCR plate to make a final reaction volume of 25 µl. We then carried out reverse transcription (RT) for 30 minutes at 50°C, followed by RT inactivation for 2 minutes at 95°C, fluorescence detection for 15 seconds at 95°C, and annealing for 1 minute at 60°C. Additionally, the assay includes an internal control reaction targeting human endogenous ribonuclease P (RP) to ensure the extracted test specimen contains amplifiable RNA. The fluorescent signal intensity was captured using a real-time PCR instrument called QuantStudio5 Real-Time PCR system (ThermoFisher Scientific).

### Dengue serotyping

Following PCR screening, samples positive for DENV were serotyped from the same RNA extract using CDC DENV 1-4 rRT-PCR Multiplex assay on the QuantStudio5 Real-Time PCR system. The assay employed DENV 1-4 specific primers and probes provided by the CDC, USA. To perform the assay, we mixed 5 µl of extracted RNA templates with 12.5 µl of the master mix (SuperScript™ III One-Step RT-PCR System with Platinum™ Taq DNA Polymerase, ThermoFischer Scientific). The assay was run in a multiplex format, with each serotype-specific probe labeled with different probes as per the manufacturer’s protocol.

### Dengue sequencing

After serotyping, we selected specimens with PCR screening Ct values below 26 for sequencing. Sequencing was carried out with technical support from CERI and KRISP. The primer scheme for DENV sequencing and other arboviruses is available on the CLIMADE GitHub (https://github.com/CERI-KRISP/CLIMADE/tree/master/Protocols/Arboviruses). The COVIDSeq protocol was adapted to perform the library preparation, followed by sequencing on the Illumina Miseq platform. This protocol is available on the CLIMADE GitHub website, and it is described in detail by *Elaine Vieira Santos, Debora Glenda Lima de La Roque in protocols.io (*https://protocols.io/view/pathogen-whole-genome-sequencing-multiplexed-ampli-cgwbtxan).

The libraries were prepared using the Illumina COVIDSeq protocol (Illumina Inc, USA). Briefly, RNA was reverse transcribed to cDNA using random hexamers. The Dengue genome was amplified using the two pools of primers specific for the Dengue serotype 1-4. PCR amplicons were tagmented using the EBLTS (Enrichment BLT), which is a process that fragments and tags the PCR amplicons with adapter sequences. Adaptor ligated amplicons were further amplified using the unique 10 base pair Index 1 (i7) adapters and Index 2 (i5) adapters (IDT for Illumina-PCR Indexes Set 1) for each sample. The pooled amplicon library was quantified using a Qubit 2.0 fluorometer (Invitrogen, Inc.) and diluted to a 4 nM. The 4nM library was denatured and diluted to a final loading concentration of 12pM. Dual indexed paired-end sequencing was performed on the Illumina Miseq using a v3 600 cycle flow cell.

### Bioinformatics analysis

Following base calling and demultiplexing of the sequence run, fastq files were processed using the Genome Detective analysis pipeline version 2.13.3 (https://www.genomedetective.com/). Upon completion, consensus fasta and Binary Alignment Map (BAM) files were retrieved from Genome Detective for each individual sample. Sequence genotyping and lineage classification was performed using the Genome Detective Dengue Typing Tool (https://www.genomedetective.com/app/typingtool/dengue/), based on a newly developed nomenclature for DENV classification (20). The resulting genomic sequences have been made available on the GISAID Epi-Arbo database (EPI_ISL_19229161 - EPI_ISL_19229193).

### Sequence Alignment and Phylogenetic Analysis

DENV reference datasets were retrieved from two primary databases: the National Centre for Biotechnology Information (NCBI) GenBank and the GISAID Epi-Arbo web interface (https://gisaid.org/). Any duplicate entries were removed. Sequence genotyping and lineage classification was performed using the Genome Detective Dengue Typing Tool (https://www.genomedetective.com/app/typingtool/dengue/), based on a newly developed nomenclature for DENV classification (20). The retrieved sequences corresponding to the lineages detected from newly sequenced Ethiopian genomes were subjected to initial quality control to remove any unverified sequences and incomplete records (i.e. geographical location and sampling dates). The final dataset used for the phylogenetic analyses in this study consisted of 33 sequences generated in this study from Ethiopia, as well as 2348 publicly available sequences of DENV3 genotype III major lineage B (corresponding to one of the transmission lineages detected in Ethiopia) and 990 sequences of DENV1 genotype III major lineage A (corresponding to the other transmission lineage detected in Ethiopia). Our DENV1 and DENV3 datasets were aligned against the appropriate DENV serotype reference genome (DENV1: NC_001477.1; DENV3: NC_001475.2) using Nextalign v1.3.0 alignment tool (21).

Maximum likelihood trees for each of the two genotypes were generated using IQ-TREE v 2.2.2.2 with 1,000 bootstraps (22). The nucleotide substitution model TIM2+F+I+G4 was selected for DENV1 and GTR+F+I+G4 for DENV3, respectively, according to Bayesian Information Criterion using Model Finder (23). The molecular clock signal was evaluated using TempEst v1.5.3 (24). Potential outliers that violated the molecular clock assumption were removed prior to inferring a time-scaled tree (TreeTime v 0.10.0) using a molecular clock rate of 5.015 x10^-4^ and 1.225×10^-4^ for DENV1 and DENV3 respectively nucleotide substitutions per site per year, as determined by the clock function in TreeTime.

### Time-calibrated Bayesian phylogenies

Time-calibrated Bayesian phylogenetic trees were constructed to estimate the time of emergence (time to the most common recent ancestor, TMRCA) of the transmission lineages circulating during the 2023 outbreak, and the likely introduction routes. The latest release of BEAST (v.1.10.4) was used along with the BEAGLE library v3.2.0 to improve computational speed (25, 26). A relaxed clock model was applied for all analyses along with the HKY+G+I nucleotide substitution model. A constant population coalescent model assumption was used. Subsequently, all analyses were performed in two independent runs of 100 million iterations each. Convergence of MCMC chains was checked using Tracer v.1.7.1 (27) and 10% of initial chains was discarded as burn-in. Post-burn-in samples were pooled to summarize parameter estimates using LogCombiner and TreeAnnotator, including posterior probability of each parameter and maximum clade credibility (MCC) trees.

### Visualisations

The phylogenetic trees were visualized using Figtree v1.4.4 (http://tree.bio.ed.ac.uk/software/figtree/) and other figures using R and ggplot (28).

### Air Travel Data

We evaluated travel data generated from the International Air Transport Association (IATA) to quantify passenger volumes originating from international airports into. IATA data accounts for ∼90% of passenger travel itineraries on commercial flights, excluding transportation via unscheduled charter flights (the remainder is modeled using market intelligence).

### Ethical considerations

The study obtained ethical approval from Addis Ababa University (Ref. no. CNCSDO/175/15/2023), and Institutional Review Boards (IRBs) of the EPHI (EPHI-IRB-536-2023).

## Results

Participants were recruited during a recorded DENV outbreak in Ethiopia (Supplementary Figure 1). Cases were recorded in multiple regions of the country by the Ministry of Health (MoH) in Ethiopia from early 2023, with Afar and Dire Dawa being the two most affected regions (Figure 1A, C). There was a first peak in cases in the Afar region between April and August 2023, low level transmission in Dire Dawa over that period, and a second prominent peak in Dire Dawa from October 2023 to January 2024. Our clinical surveillance was able to detect dengue acute infections from both these regions, with detection in Dire Dawa intensifying prior to the peak in recorded cases (Figure 1A).

**Figure 1.**
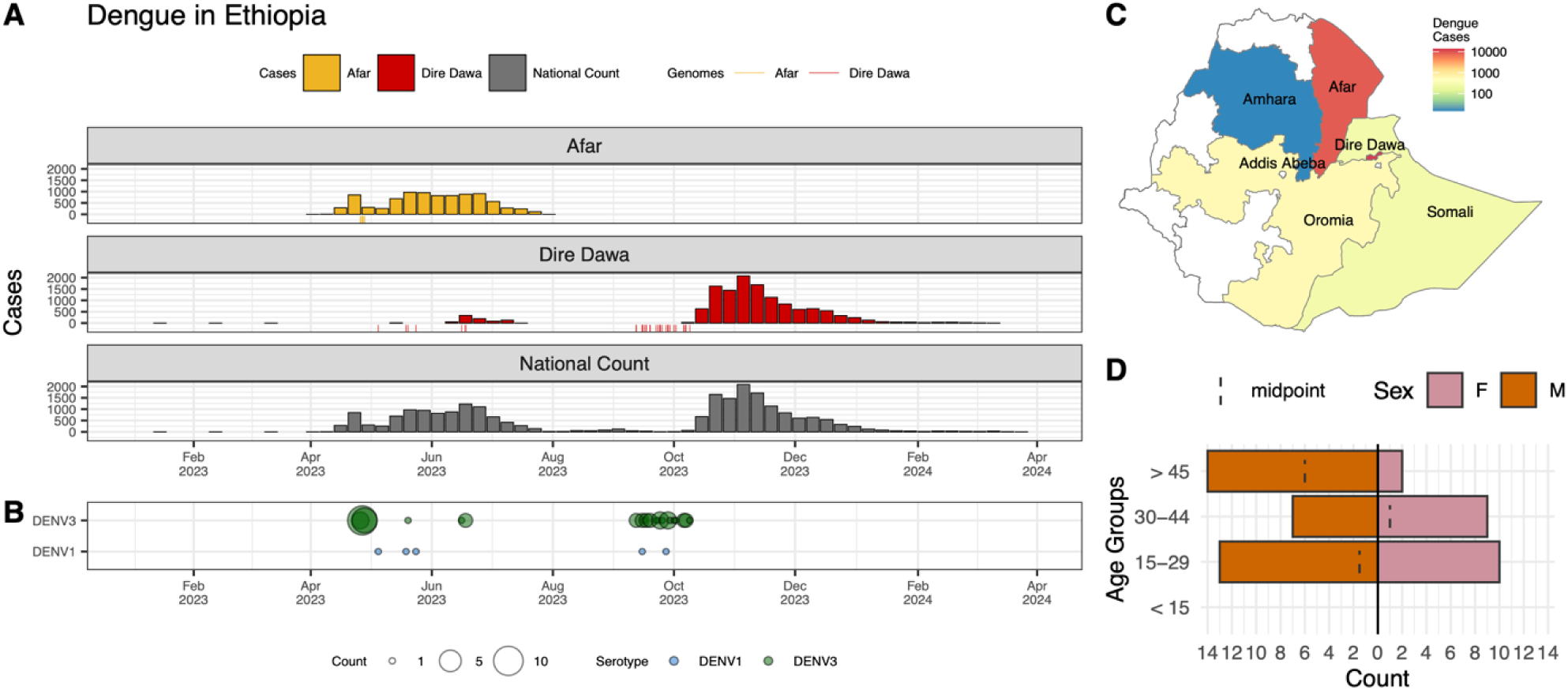
Spatiotemporal distribution of dengue epidemiology and ecology in Ethiopia. A) Epidemiological curve of dengue cases in Ethiopia: Afar region (outbreak from April 2023 to August 2023), Dire Dawa City Administration (outbreak from June 2023 to April 2024), and national count from 2023 to 2024. B) Serotype distribution of sampled cases. C) Map of DENV cases in Ethiopia from 2023 to 2024, with the Afar region and Dire Dawa City administration outlined in red. D) Demographic distribution of sampled cases.

### Sample collection and dengue epidemiology in Ethiopia

Out of 891 febrile patients screened for DENV, CHIKV, and ZIKV infections, only dengue virus was detected in this study. The overall proportion with a positive PCR for DENV was 10.4% (93 out of 891 patients). The test positivity was slightly higher in men than women (Table 1). DENV was isolated from two of the sites; the proportion with a positive DENV PCR was 13.7% (41/300) in Afar and 17.9% (52/291) in Dire Dawa. None of the 300 samples from Gambela were positive for DENV. Virus genotyping shows evidence of co-circulation of DENV1 and DENV3 throughout the outbreak period (Figure 1B). Of the 93 isolates, 88 (94.6%) were identified as belonging to the DENV3 serotype, while the remaining 5 (5.4%) were identified as DENV1. At the Afar site, only DENV3 was detected (n=36), whereas at the Dire Dawa site, both DENV1 (n=5) and DENV3 (n=47) were detected (Table 1).

**Table 1:**
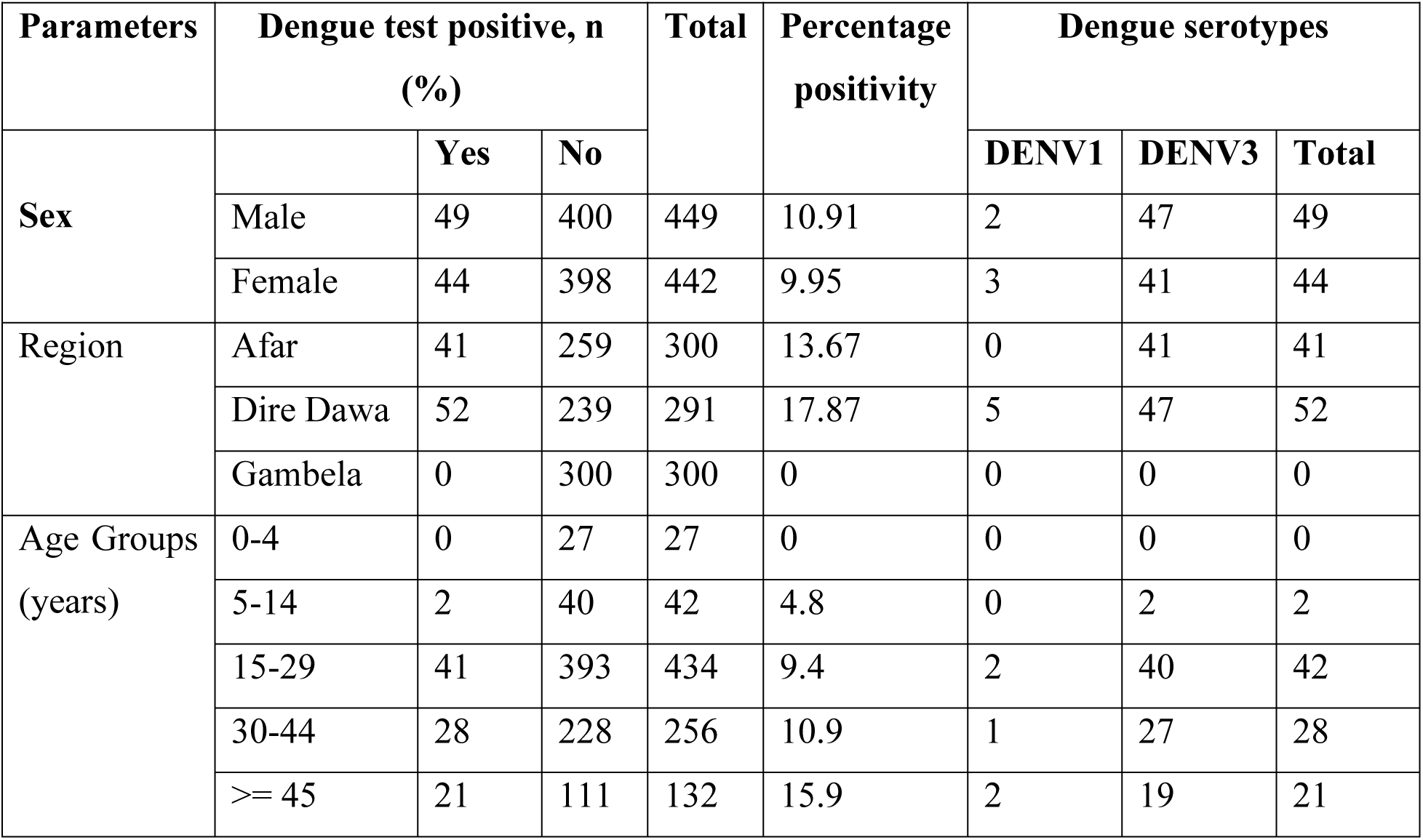
Socio-demographics characteristics, burden of DENV infection, and serotype circulation in Ethiopia.

| Parameters | Dengue test positive, n (%) |  |  | Total | Percentage positivity | Dengue serotypes |  |  |
| --- | --- | --- | --- | --- | --- | --- | --- | --- |
|  |  | Yes | No |  |  | DENV1 | DENV3 | Total |
| Sex | Male | 49 | 400 | 449 | 10.91 | 2 | 47 | 49 |
|  | Female | 44 | 398 | 442 | 9.95 | 3 | 41 | 44 |
| Region | Afar | 41 | 259 | 300 | 13.67 | 0 | 41 | 41 |
|  | Dire Dawa | 52 | 239 | 291 | 17.87 | 5 | 47 | 52 |
|  | Gambela | 0 | 300 | 300 | 0 | 0 | 0 | 0 |
| Age Groups (years) | 0-4 | 0 | 27 | 27 | 0 | 0 | 0 | 0 |
|  | 5-14 | 2 | 40 | 42 | 4.8 | 0 | 2 | 2 |
|  | 15-29 | 41 | 393 | 434 | 9.4 | 2 | 40 | 42 |
|  | 30-44 | 28 | 228 | 256 | 10.9 | 1 | 27 | 28 |
|  | >= 45 | 21 | 111 | 132 | 15.9 | 2 | 19 | 21 |

### Virus genome sequencing

A total of 55 viral isolates were selected based on their PCR screening Ct values from Afar region and Dire Dawa City administration. The Ct values of the isolates ranged from 15.5 to 25.6. A total of 20 DENV3 were selected from Afar and 35 (5 DENV1 and 30 DENV3) were selected from Dire Dawa sites. Out of these, 54 isolates were successfully sequenced, and only one sample failed to be sequenced due to low sample volume. Among the successfully sequenced isolates, 33 gave near whole genome sequences (>90% genome coverage). Of these, all of the 5 DENV1 identified as DENV1 genotype III major lineage A (DENV1_IIIA), while the remaining 28 were identified as DENV3 genotype III major lineage B (DENV3_IIIB), with 1 isolate from Afar and 27 from Dire Dawa (Figure 2).

**Figure 2.**
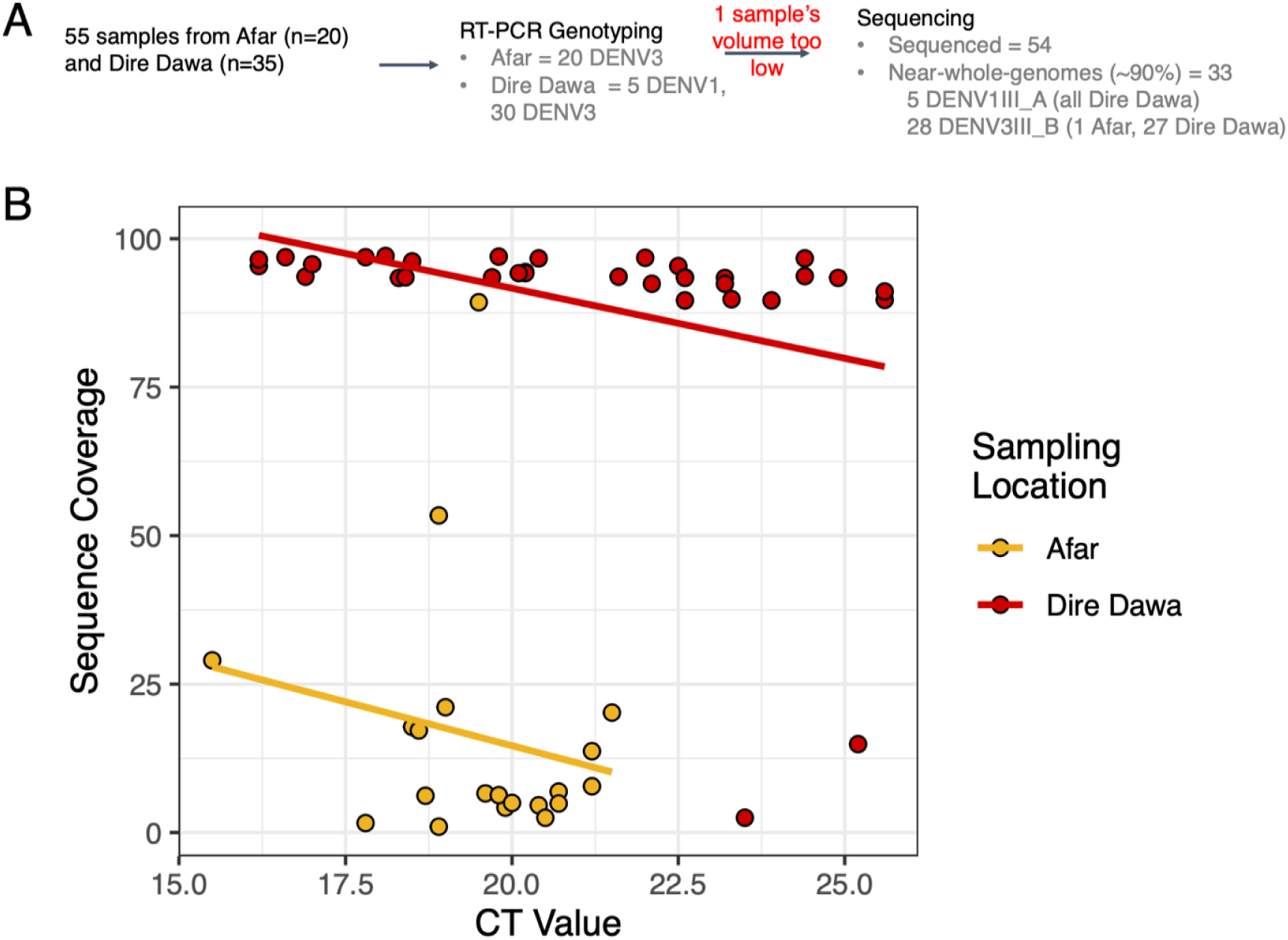
Sequencing process and coverage results. A) Sample selection, genotyping and sequencing workflow. B) CT values versus sequence coverage for all sequenced specimens.

Samples collected from Dire Dawa demonstrated an expected association between a high viral load (lower Ct values) and high sequence coverage whereas most samples from Afar resulted in low sequence coverage genomes despite low Ct values. The Ct values were not repeated after storing the serum samples; we only used the first screening PCR Ct values. The sample collection and storage freezers used were different for both sites. The samples from Afar site were collected in DNA/RNA shield during the outbreak in May 2023 and stored for a long time at −20°C, with intermittent temperature fluctuations of the freezer. In contrast, samples collected from Dire Dawa and Gambela sites were relatively recent and stored at −80°C until they were sequenced.

### Evolutionary history of sequences belonging to Ethiopia’s 2023 outbreak

A phylogenetic tree was reconstructed with 990 global DENV1_IIIA aligned sequences, including five from Ethiopia sequenced in this study (Figure 3A). For this lineage, all sequences from Ethiopia belonged to a single transmission cluster, which clustered monophyletically, with other African sequences (Democratic Republic of Congo and Djibouti), suggesting cryptic transmission in the region since 2019 (Figure 3B). TMRCA of the Ethiopian clade specifically to mid 2021 (Figure 3B). Basal to this African clade are sequences mostly of Asian origin suggested an introduction into Africa from Asia, which we can date back to the year 2018 (Figure 3B). For the DENV3_IIIB lineage, we constructed a phylogenetic tree with 2,348 global sequences, including the ones from Ethiopia (Figure 3C). Sequences from Ethiopia belong to two distinct clades on the tree (Figure 3D, E). While the first clade consisted of sequences generally of Asian and African origin, the Ethiopian sequences, with a TMRCA of early 2021 (95% HPD: 2020 – 2022) cluster monophyletically with a sequence from Italy’s 2023 outbreak (29). We infer that the common ancestor between the Ethiopian isolates and the Italy one existed around mid-2019 (95% HPD: mid-2017 - 2021) with long branches separating the two locations. The Ethiopia-Italy clade is supported with >70% posterior support at the relevant internal nodes on the MCC tree. This suggests some level of cryptic transmission in unsampled areas from where both Ethiopia and Italy could have received viral introductions. However, given historical ties between the two countries and the remaining high connectivity, Italy being the European country with the second most air travel passengers into Ethiopia (Supplementary Figure 2), an introduction from there is plausible. The second relevant clade of this lineage shows a clear introduction from Asia to Ethiopia late 2021 (95% HPD: 2021 - 2023), with potential persistent transmission since then (Figure 3E). While the closest relatives to this Ethiopian clade are sequences from India, it remains possible, again given long branch lengths on the tree, that intermediate unsampled regions are involved, including country such as Saudi Arabia (30), which is the top countries with largest passenger volume to Ethiopia (Supplementary Figure 2).

**Figure 3.**
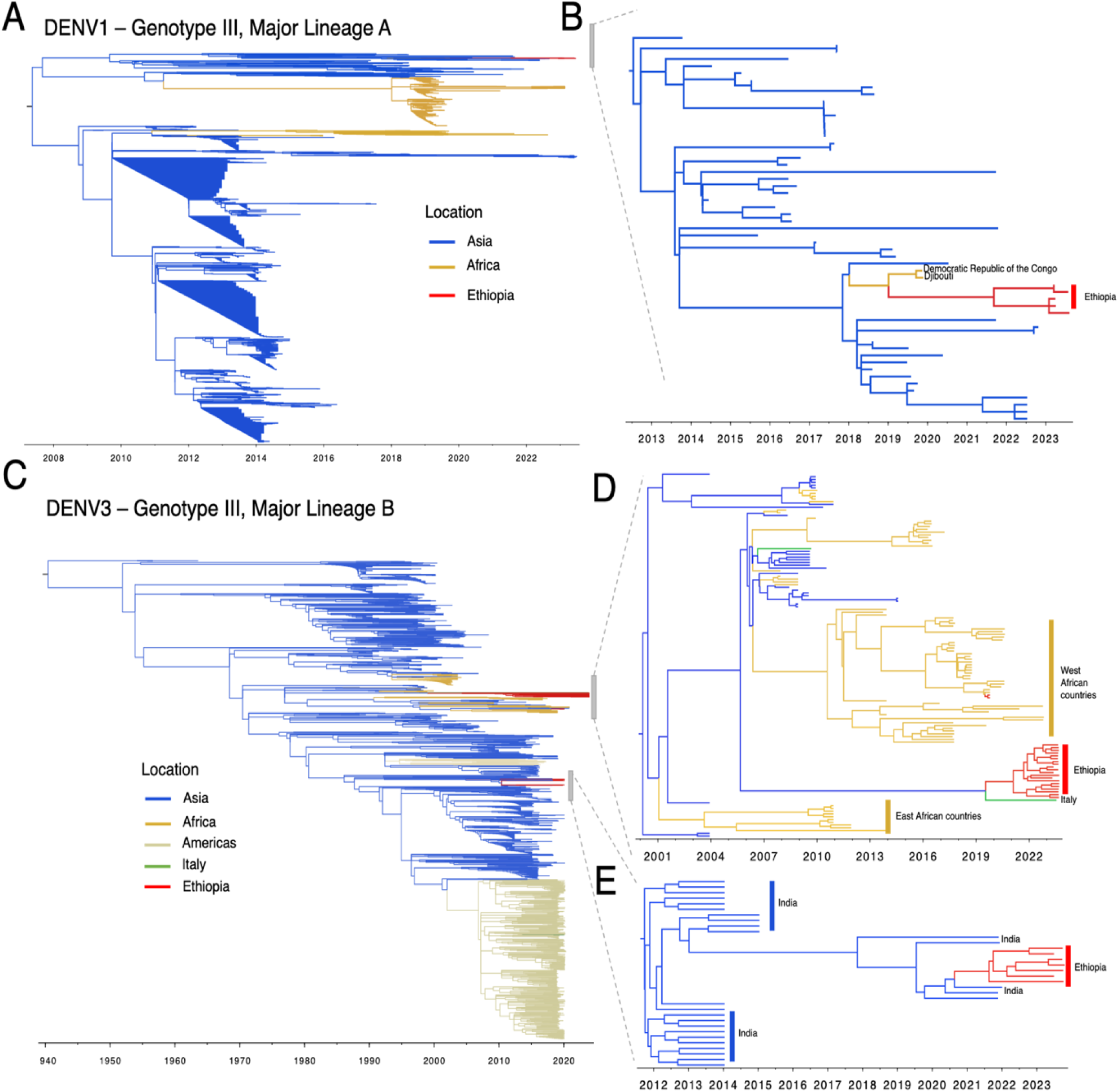
Phylogenetic analysis of Ethiopian genomes. A) Timed maximum likelihood phylogeny of the DENV1 genotype III major lineage A clade, containing Ethiopian genomes sequenced in this study. The tree is coloured by sequence locations. B) Zoom-in of a sub-clade of the tree to show close evolutionary relationships related to the Ethiopian DENV1 genomes. C) Timed maximum likelihood phylogeny of the DENV3 genotype III major lineage B clade, containing Ethiopian genomes sequenced in this study. The tree is coloured by sequence locations. D) Maximum clade credibility (MCC) tree of sub-tree containing cluster 1 of Ethiopian DENV3 genomes. E) Maximum clade credibility (MCC) tree of sub-tree containing cluster 2 of Ethiopian DENV3 genomes.

## Discussion

In this study, we investigated the genomic epidemiology of DENV in two areas of Ethiopia. This study presents the distribution of serotypes and first-ever report of the genotypes and lineages circulating in Ethiopia from the first ever whole genomes sequenced in the country.

Our analysis showed that two serotypes, DENV1 and DENV3 were isolated from Dire Dawa, while only DENV3 was isolated in the Afar area. These preliminary data suggest a need for more comprehensive epidemiological or genomic surveillance and strengthened systems for severe dengue surveillance due to the potential risk of severe dengue from multiple serotypes or genotypes. In Africa, all four serotypes of DENV have been detected in both humans and *Aedes* mosquitoes (31). DENV2 is the most prevalent serotype in the East African region. The virus caused outbreaks in Ethiopia in 2013, Kenya in 2013, Tanzania in 2014, and Mozambique in 2014-2015, and has continued to remain prevalent in these areas (32–34). DENV1 outbreaks have been detected at different times in Angola, Kenya, Senegal, and Somalia (35). The current DENV1 serotype may have been historically co-circulating with DENV2 or may have been a recent introduction into Dire Dawa.

The current outbreak of dengue fever first began in the Mile district of the Afar Region in Ethiopia in April 2023, in which the outbreak was caused by DENV3 and has since spread to at least five other regions in the Eastern part of the country (36). Between April 2, 2023, and February 26, 2024, dengue cases were reported to the Public Health Emergency Operations Center in Ethiopia from these affected regions.

Phylogenetic analysis revealed that the DENV1 serotype belongs to genotype III major lineage A. The analysis also showed that the circulation of this lineage during the 2023 outbreak corresponds to a single transmission cluster. It seems that this cluster was introduced to Ethiopia from Asia via other African countries. Dengue viruses belonging to the same genotype were sequenced during the 2023 dengue outbreak in Chad (2). However, the Ethiopian genomes generated in this study are not directly linked to the outbreak in Chad, which originated from a large outbreak in Tanzania in 2019, via Nigeria (37). The phylogenetic reconstruction from available DENV1 genomic data in Africa, which we have supplemented with our sequences, show that several distinct lineages are currently circulating on the continent. All known Central African DENV1 strains belong to the genotype V African lineage, with the oldest strains of this lineage being isolated in Nigeria in 1968 (38). However, it remains that the lack of publicly available data limits our understanding of the dynamics of DENV1 within the continent.

On the other hand, the DENV3 viruses sequenced during Ethiopia’s 2023 outbreak belonged to the genotype III major lineage B, and phylogenetic reconstructions revealed two transmission clusters circulating in Ethiopia. Cluster-1 is closely related to a DENV3 virus sequenced in 2023 sequence from Italy (as shown in Figure 3A) (29). This highlights the possibility that the virus may have been introduced into Ethiopia from Italy. Since we have no previous sequence information, we cannot exclude the possibility of transmission from Ethiopia to Italy.

Available data indicates that DENV3 genotype III exists in neighboring countries such as Sudan, Kenya, Djibouti, and Somalia (39). However, the current DENV3 genotype III in Ethiopia is not directly related to the African isolates. Cluster-2 appears to have been introduced from India perhaps via secondary locations (as shown in Figure 3E). Phylogenetic analysis of DENV3 in India has shown that outbreaks were caused by genotype III and the outbreak during 2017-2018 was similar to the strain that was circulating in 2016 (40).

While performing the whole genome sequencing, only 33 samples resulted in approximately 90% and above genome coverage. Thirty-two (DENV1 =5 and DENV3 =27) of these samples were from Dire Dawa site but only one sample gave approximately 90% genome coverage from Afar site. Serum samples from the Afar site was collected during the outbreak in tubes containing DNA/RNA shield solution and sent to the Ethiopian Public Health Laboratory for detection and serotyping. The remaining samples were then stored at −20°C for around 11 months before sequencing. Sequencing was carried out following this period of storage. The limited genome coverage observed could be attributed to the extended storage under unsuitable conditions. In contrast, serum samples obtained from Dire Dawa were gathered without DNA/RNA shield and were relatively recent, stored at −80°C for approximately 4 to 8 months, resulting in greater sequence coverage.

This study presents the first report on the genomic epidemiology of the 2023 dengue outbreak in Ethiopia. However, it is important to note some limitations: 1) Poor sequencing outcomes were encountered, possibly due to inadequate sample storage at Afar. Improved sample storage conditions could have led to better whole genome sequencing results. 2) The limited availability of DENV sequences continentally and globally restricts our ability to infer global transmission dynamics and direct introduction routes. In this study, we investigated the cocirculation of DENV1 and DENV3 in Ethiopia during the 2023 outbreak. We were able to identify circulation genotypes and lineages and infer possible introduction routes and timings. Our findings highlight the utility for comprehensive disease surveillance, including pathogen genome sequencing, of dengue transmission in Ethiopia and beyond to understand transmission dynamics of emerging outbreaks. Regular active surveillance through highly connected sentinel sites as well as at port of entry can improve response time to outbreaks and reduce the likelihood of dengue establishment.

## Supplementary Materials

**Supplementary Figure 1:**
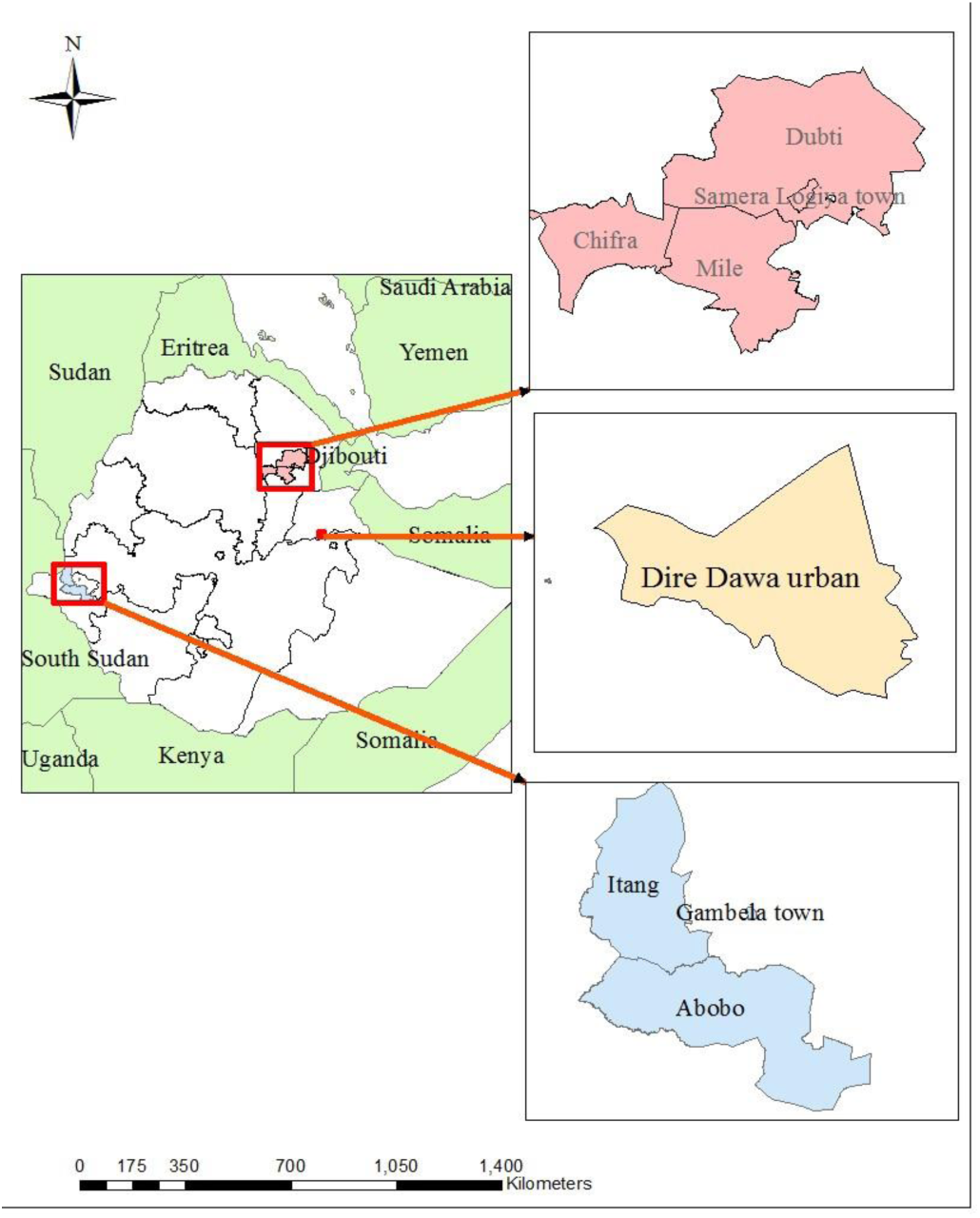
Map showing the location of hospital-based sentinel sites where samples were collected for the study. The Afar region and Dire Dawa City administration are located in the eastern part of Ethiopia, while the Gambela region is situated in the western part of Ethiopia.

**Supplementary Figure 2:**
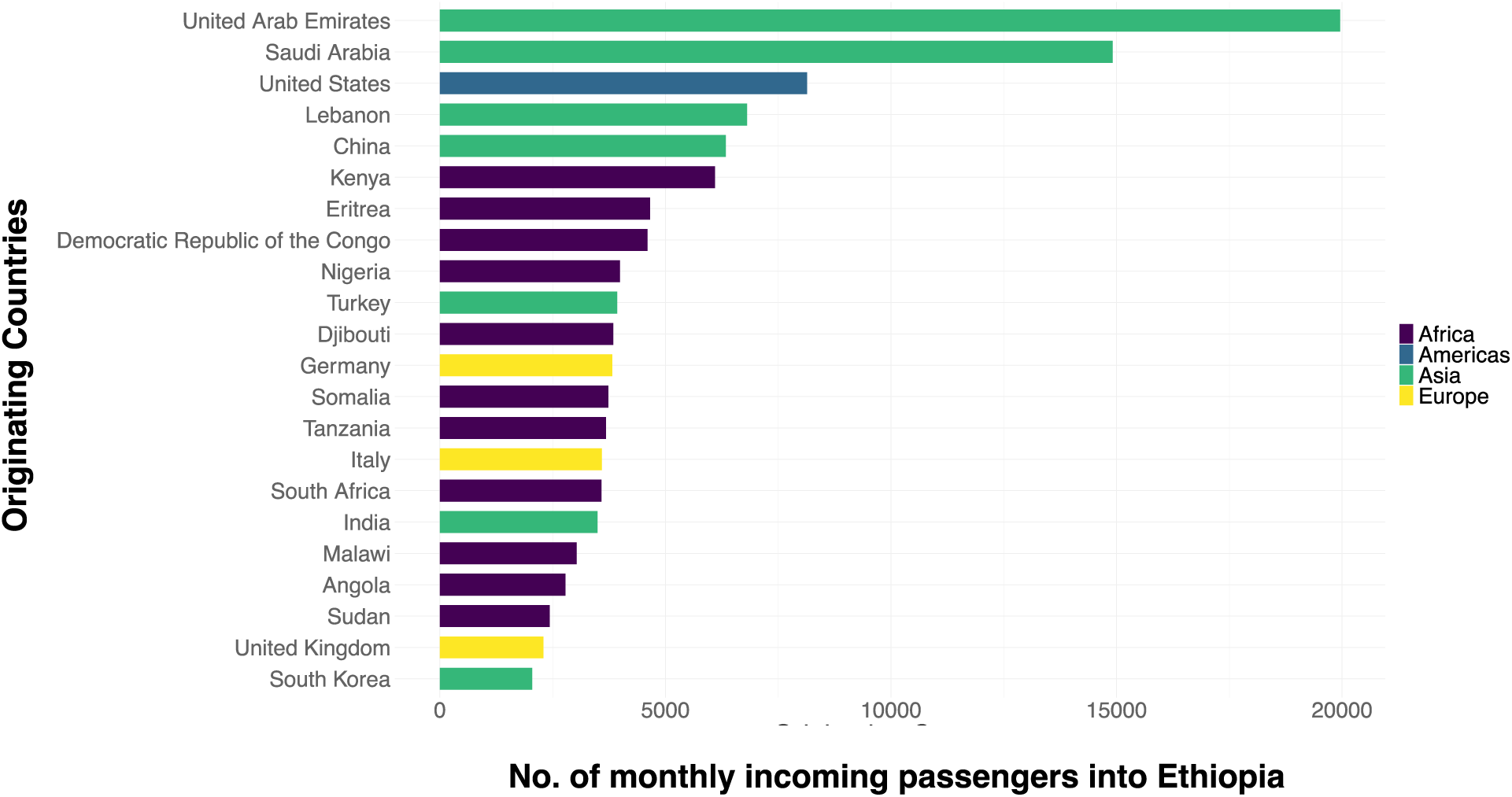
Monthly incoming travel into Ethiopia from countries with >2000 incoming passengers. The data shown is for February 2020 as a representative month.

**Supplementary Table 1:** Acknowledgement of Authors for data obtained from the GISAID EpiArbo database.

## Author Contributions

Ad.A. and D.B. conceived and designed the experiments; AdA., H.T. and E.W. performed the data analysis and experiments; Ab.A., F.R., M.B., H.T., K.K., C.H., M.U.G.K, T.d.O. contributed materials/analysis tools; J.G., and S.P. participated in the wet laboratory work; Ge.Ta., Ge.To., D.B., R.de.W.T., T.d.O., D.W., supervised the work; and Ad.A., H.T. E.W., and D.B. wrote the original manuscript and the paper. All authors have read and agreed to the published version of the manuscript.

## Informed Consent Statement

All participants gave written informed consent for participation and publication.

## Data Availability Statement

All sequences used for this manuscript are publicly available on the GISAID EpiArbo database, subject to its terms and conditions (https://www.gisaid.org/), GISAID identifier: EPI_ISL_19229161 - EPI_ISL_19229193.

## Acknowledgments

Concerning our reference data against which we analysed the genomes generated in this study, where a portion of those were obtained from the GISAID EpiArbo database, we gratefully acknowledge Authors from the Originating laboratories responsible for obtaining the specimens, as well as the Submitting laboratories where the genome data were generated and shared via GISAID (See Supplementary Table 1). Additionally, we extend our gratitude to CDC Atlanta for supplying Dengue detection and serotyping kits.

## Abbreviations

BAM: Binary Alignment Map
CDC: Center for Diseases Control and prevention
CERI: Center for Emerging and Re-emerging Infectious diseases
CHIKV: Chikungunya virus
CLIMADE: Climate Amplified Diseases and Epidemics
COVID-19: Corona Virus Diseases-19
DENV: Dengue virus
EPHI: Ethiopian Public Health Institute
GISAID: Global initiative on sharing all influenza data
HPD: Highest Posterior Density
IATA: International Air Transport Association
IRB: Institutional Review Board
KRISP: KwaZulu-Natal Research Innovation and Sequencing Platform
MCC: Maximum Clade Credibility
NCBI: National Centre for Biotechnology Information
RT-PCR: Reverse transcription polymerase chain reaction
TMRCA: The Most Common Recent Ancestor
ZIKV: Zika virus

## References

1. World Health Organization. Disease Outbreak News; Dengue – Global situation. 2023. Available at: https://www.who.int/emergencies/disease-outbreak-news/item/2023-DON498.

2. WHO African Region Health Emergency Situation Report-Multi-country Outbreak of Dengue, Consolidated Regional Situation Report # 001. 2023.

3. Ethiopia Steps Up Actions for Dengue Prevention and Control. World Health Organization Africa. 2014. Available from: https://www.afro.who.int/news/ethiopia-steps-actionsdengue-prevention-and-control.

4. Ahmed YM, Salah A.A. Epidemiology of Dengue Fever in Ethiopian Somali Region: Retrospective Health Facility Based Study. Central African Journal of Public Health. 2016; 2(2): 51–56. doi: 10.11648/j.cajph.20160202.12.

5. Gutu MA, Bekele A, Seid Y, Mohammed Y, Gemechu F, Woyessa AB, et al. Anotherdengue fever outbreak in Eastern Ethiopia—An emerging public health threat. PLoS Negl Trop Dis 2021; 15(1): e0008992. 10.1371/journal.pntd.0008992.

6. Hasan M.M, Hernandez-Y P.J., Rivera-Cabrera M.A., Sarkar A., Santos Costa A.C., Essar M.Y. Concurrent epidemics of dengue and COVID-19 in Peru: Which way forward? The Lancet Regional Health – Americas. 2022;12: 100277. 10.1016/j.lana.2022.100277.

7. Andhikaputra, G., Lin, YH., Wang, YC. Effects of temperature, rainfall, and El Niño Southern Oscillations on dengue-like-illness incidence in Solomon Islands. BMC Infect Dis. 2023; 23: 206. 10.1186/s12879-023-08188-x.

8. Murugesan A, Manoharan M. Dengue Virus. In book of Emerging and Reemerging Viral Pathogens. Chapter 2. 2020: 281–359. 10.1016/B978-0-12-819400-3.00016-8.

9. Mustafa M.S., Rasotgi V., Jain S., Gupta V. Discovery of fifth serotype of dengue virus (DENV-5): A new public health dilemma in dengue control. Medical Journal Armed Forces India. 2015; 71(1): 67–70. 10.1016/j.mjafi.2014.09.011.

10. Poltep K., Phadungsombat J., Nakayama E.E., Kosoltanapiwat N., Hanboonkunupakarn B., Wiriyarat W., Shioda, T., Leaungwutiwong P., Genetic Diversity of Dengue Virus in Clinical Specimens from Bangkok, Thailand, during 2018–2020: Co-Circulation of All Four Serotypes with Multiple Genotypes and/or Clades. Trop. Med. Infect. Dis. 2021; 6:162. 10.3390/tropicalmed6030162.

11. Calvez E, Somlor S, Viengphouthong S, Balière C, Bounmany P, Keosenhom S, et al. Rapid genotyping protocol to improve dengue virus serotype 2 survey in Lao PDR. PLoS ONE. 2020; 15(8): e0237384. 10.1371/journal.pone.0237384.

12. Li L., Guo X., Zhang X., Lingzhai Zhao L., Li L., Wang Y., et al. A unified global genotyping framework of dengue virus serotype-1 for a stratified coordinated surveillance strategy of dengue epidemics. Infect Dis Poverty. 2022; 11:107. 10.1186/s40249-022-01024-5.

13. Naveca F., Santiago G. A., Maito R., Ribeiro Meneses C., do Nascimento V., de Souza V., Bello G. Reemergence of Dengue Virus Serotype 3, Brazil, 2023. Emerging Infectious Diseases. 2023; 29(7):1482–1484. 10.3201/eid2907.230595.

14. Hill V, Cleemput S, Fonseca V, Tegally H, Brito AF, Gifford R, et al,. A new lineage nomenclature to aid genomic surveillance of dengue virus. medRxiv [Preprint]. 2024 May 17:2024.05.16.24307504. doi: 10.1101/2024.05.16.24307504.

15. Rahim R, Hasan A, Phadungsombat J, Hasan N, Ara N, Biswas SM, Nakayama EE, Rahman M, Shioda T. Genetic Analysis of Dengue Virus in Severe and Non-Severe Cases in Dhaka, Bangladesh, in 2018-2022. Viruses. 2023;15(5):1144. doi: 10.3390/v15051144.

16. Ahamed S.F., Rosario V., Britto C., Dias M., Nayak K., Chandele A., Kaja M-K., Shet A. Emergence of new genotypes and lineages of dengue viruses during the 2012–15 epidemics in southern India. International Journal of Infectious Diseases. 2019; 84 (2019): S34–S43. 10.1016/j.ijid.2019.01.014.

17. Durbin AP, Whitehead SS. Next-Generation Dengue Vaccines: Novel Strategies Currently Under Development. Viruses. 2011; 3(10):1800–1814. 10.3390/v3101800.

18. Asebe G, Michlmayr D, Mamo G, Abegaz WE, Endale A, Medhin G, et al. Seroprevalence of Yellow fever, Chikungunya, and Zika virus at a community level in the Gambella Region, South West Ethiopia. PLoS ONE. 2021; 16(7): e0253953. 10.1371/journal.pone.0253953.

19. Horstick O., Martinez E., Guzman M. G., Martin J. L., Ranzinger S. R. WHO dengue case classification 2009 and its usefulness in practice: an expert consensus in the Americas. Pathogens and global health. 2015; 109(1), 19–25. 10.1179/2047773215Y.0000000003.

20. Hill V, Cleemput S, Fonseca V, Tegally H, Brito AF, Gifford R, et al,. A new lineage nomenclature to aid genomic surveillance of dengue virus. medRxiv [Preprint]. 2024. doi: 10.1101/2024.05.16.24307504.

21. Aksamentov I., Roemer C., Hodcroft E., Neher R. Nextclade: clade assignment, mutation calling and quality control for viral genomes. J Open Source Softw. 2021; 6:3773.

22. Nguyen L.T, Schmidt HA, von Haeseler A, Minh B. Q. IQ-TREE: a fast and effective stochastic algorithm for estimating maximum-likelihood phylogenies. Mol. Biol. Evol. 2015; 32: 268–274.

23. Rambaut A, Lam TT, Max Carvalho L, Pybus OG. Exploring the temporal structure of heterochronous sequences using TempEst (formerly Path-O-Gen). Virus Evol. 2016; 2: vew007.

24. Sagulenko P, Puller V, Neher RA. TreeTime: maximum-likelihood phylodynamic analysis. Virus Evol.2018; 4: vex042.

25. Suchard MA, Lemey P, Baele G, Ayres DL, Drummond AJ, Rambaut A. Bayesian phylogenetic and phylodynamic data integration using BEAST 1.10. Virus Evol. 2018; 8;4(1): vey016. doi: 10.1093/ve/vey016.

26. Ayres DL, Darling A, Zwickl DJ, Beerli P, Holder MT, Lewis PO, et al. BEAGLE: An Application Programming Interface and High-Performance Computing Library for Statistical Phylogenetics. Syst. Biol. 2012; 61(1):170–173. 10.1093/sysbio/syr100.

27. Rambaut A, Drummond AJ, Xie D, Baele G, Suchard MA. Posterior summarization in Bayesian phylogenetics using Tracer 1.7. Syst. Biol. 2018; 67: 901–904.

28. Wickham H. ggplot2: Elegant Graphics for Data Analysis. Springer New York, NY. 2016. 10.1007/978-0-387-98141-3.

29. Branda F, Nakase T, Maruotti A, Scarpa F, Ciccozzi A, Romano C, et al., Dengue virus transmission in Italy: surveillance and epidemiological trends up to 2023. medRxiv. 2023; 12:1923300208; doi: 10.1101/2023.12.19.23300208.

30. Hashem AM, Sohrab SS, El-Kafrawy SA, Abd-Alla AMM, El-Ela SA, Abujamel TS, et al., Diversity of dengue virus-3 genotype III in Jeddah, Saudi Arabia. Acta Trop. 2018;183:114–118. doi: 10.1016/j.actatropica.2018.04.002.

31. Amarasinghe A, Kuritsk JN, Letson GW, Margolis HS. Dengue virus infection in Africa. Emerg Infect Dis. 2011;17(8):1349–54. doi: 10.3201/eid1708.101515.

32. Woyessa AB, Mengesha M, Kassa W, Kifle E, Wondabeku M, Girmay A, Kebede A. et al. The first acute febrile illness investigation associated with dengue fever in Ethiopia, 2013: A descriptive analysis. Ethiop. J. Health Dev. 2014;28(3):155–161.

33. Ellis EM, Neatherlin JC, Delorey M, Ochieng M, Mohamed AH, Mogeni DO, et al. A Household Serosurvey to Estimate the Magnitude of a Dengue Outbreak in Mombasa, Kenya, 2013. PLoS Negl Trop Dis. 2015; 9(4): e0003733. 10.1371/journal.pntd.0003733.

34. Mboera LEG, Mweya CN, Rumisha SF, Tungu PK, Stanley G, Makange MR, et al. The Risk of Dengue Virus Transmission in Dar es Salaam, Tanzania during an Epidemic Period of 2014. PLoS Negl Trop Dis. 2016; 10(1): e0004313. 10.1371/journal.pntd.0004313.

35. Diallo D., Diouf B., Gaye A., hadji Ndiaye E., Sene N.M., Dia I., et al. Dengue vectors in Africa: A review. Heliyon. 2022; 8(5): e09459. 10.1016/j.heliyon.2022.e09459.

36. Ethiopian Public Health Institute. Public Health Emergency Operations Center (PHEOC), Ethiopia. Weekly Bulletin. 2024; Bulletin number 61.

37. WHO. Weekly Bulletin on Outbreaks and Other Emergencies; Week 46. Geneva, Switzerland: WHO Regional Office for Africa; 2019.

38. Carey D.E., Causey O.R., Reddy S., Cooke A.R. Dengue viruses from febrile patients in Nigeria, 1964-68. The Lancet. 1971; 297(7690): 105–106. 10.1016/S0140-6736(71)90840-3.

39. Selhorst P., Lequime S., Dudas G., Proesmans S., Lutumba P., Katshongo F et al. Phylogeographic analysis of dengue virus serotype 1 and cosmopolitan serotype 2 in Africa. International Journal of Infectious Diseases. 2023; 133: 46–52.10.1016/j.ijid.2023.04.391.

40. Padhi A, Gupta E, Singh G, Parveen S, Islam A, Tarai B. Circulation of DENV-3 Genotype 3 during 2017 to 2018 in Delhi: A Single-Center Hospital-Based Study. J Lab Physicians. 2021; 14;14(1):21–26. doi: 10.1055/s-0041-1734017.

